# A DEFINITIVE TCRBETA1/ TCRBETA2 ANTIBODY PAIR FOR DETERMINING T-CELL MONOTYPIA AS A SURROGATE FOR CLONALITY IN LYMPHOMA DIAGNOSIS IN FORMALIN FIXED PARAFFIN EMBEDDED MATERIAL

**DOI:** 10.64898/2026.02.13.26346202

**Authors:** Anuradha Kaistha, Jinlong J. Situ, Shelley Evans, Margaret Ashton-Key, Graham Ogg, Elizabeth J. Soilleux

**Affiliations:** Department of Pathology, University of Cambridge, Tennis Court Road, Cambridge CB2 1QP, UK; Department of Cellular Pathology, University Hospital Southampton, Southampton SO16 6YD, UK; MRC Translational Immune Discovery Unit, MRC Weatherall Institute of Molecular Medicine, University of Oxford, Oxford OX3 9DS, UK

**Keywords:** T-cell lymphoma, clonality, diagnosis, TCRβ (TCRbeta), monoclonal antibody, formalin fixed paraffin embedded (FFPE), cutaneous lymphoma

## Abstract

T-cell lymphomas are often histologically indistinguishable from benign T-cell infiltrates. Clonality testing is frequently required for diagnosis. It lacks the spatial context and is slow and expensive, relying on complex, multiplexed PCR reactions, interpreted by experienced scientists or pathologists. We previously published details of a pair of highly specific monoclonal antibodies against the two alternatively used, but very similar, T-cell receptor β constant regions, TCRβ1 and TCRβ2. We demonstrated the feasibility of immunohistochemical detection of TCRβ1 and TCRβ2 in formalin-fixed, paraffin-embedded (FFPE) tissue as a novel diagnostic strategy for T-cell lymphomas. Here we validate an improved pairing of TCRβ1/2 rabbit monoclonal antibodies, and demonstrate their utility for single and double immunostaining, including with a chimeric mouse anti-TCRβ2 antibody. Finally, we show that this staining is amenable to automated cell counting, permitting accurate calculation of the TCRβ2:TCRβ1 ratio.

## Introduction

T-cell lymphomas account for 10–15% of lymphomas and generally have a rapid and aggressive course[1]. They are difficult, time-consuming and expensive to diagnose, so patients often require multiple biopsies, with months or years-long diagnostic delays impacting survival (reviewed in [1]).

T-cell lymphomas composed of small-to-medium-sized cells (e.g., peripheral T-cell lymphoma, angioimmunoblastic lymphoma, and mycosis fungoides[2]), are particularly difficult to distinguish, both morphologically and immunophenotypically, from benign lymphocytic infiltrates. Diagnosis often necessitates multiplexed PCR T-cell clonality analysis of DNA extracted from tissue, a complex test interpreted by specially trained staff members. Currently, clonality studies are only undertaken in large specialist centres and can delay diagnosis by several weeks. Compared with immunostained tissue on a slide, DNA-based testing precludes concomitant assessment of tissue architecture, morphology and immunophenotype of any cells suspected to be a clonal population[1, 3].

While clonality studies are performed on most suspected T-cell lymphomas, a much smaller proportion of suspected B-cell lymphomas or plasma cell neoplasms are tested, because B cells and plasma cells show immunoglobulin light chain restriction. This naturally occurring phenomenon permits comparison of numbers of cells expressing kappa and lambda light chains by immunohistochemistry or in situ hybridization. Light-chain monotypia (restriction to one light-chain type) acts as a surrogate for monoclonality[4-6].

Around 95% of T cells bear an αβ T-cell receptor (TCR)[2]. Gene segment rearrangement at the TCRβ (*TRB*) locus produces T cells with either a TCRβ1 or TCRβ2 constant region, encoded by *TRBC1* or *TRBC2,* respectively[7-9]. Immunohistochemical detection of the TCRβ1/2 proteins, in a manner analogous to kappa and lambda light chain detection in B cells, allows timely and cost-effective assessment of T-cell monotypia (TCRβ restriction), as a surrogate for T-cell monoclonality [1, 3].

We previously published details of an antibody pair amenable to formalin fixed paraffin embedded (FFPE) immunostaining, suitable for assessing T-cell monotypia [1]. Here we validate an improved rabbit monoclonal anti-TCRβ1/2 pairing, with a better performing anti-TCRβ2. We demonstrate their utility for single and double immunostaining, including with a chimeric mouse anti-TCRβ2 antibody. Finally, we show that this staining is amenable to automated cell counting, permitting accurate calculation of the TCRβ2:TCRβ1 ratio in each sample, and thus of benign/ malignant ratio cut-offs.

## Materials and Methods

### Production of Recombinant Rabbit and Mouse Antibodies

As described previously[1], rabbits were immunized with TCRβ1- or TCRβ2-derived peptides to produce TCRβ1-specific (ROX7) and TCRβ2-specific (ROX13) rabbit IgG antibodies. Recombinant antibodies were expressed in HEK293T cells and purified by size-exclusion HPLC (Superdex 200, GE28-9909-44, Sigma Aldrich, Gillingham, UK), to achieve >98% purity. Chimeric mouse antibody (MOX13) was produced by recombinant switching of the C-regions of the ROX13 antibody.

### Cell culture, Western blotting and the immunostaining of immortal T- and B-cell lines

Jurkat (TCRβ1-expressing), CEM, MOLT-4, and HPB-ALL (TCRβ2-expressing)[10], and Daudi (B-cell; TCRβ1/β2-negative) immortalized lymphoid cell lines were used for Western blot analysis, as previously described[1]. Briefly, cells were washed with ice-cold PBS and lysed in RIPA buffer supplemented with a protease inhibitor cocktail (cOmplete™ Mini, EDTA-free; Roche; 11836170001; 100× stock). Protein concentrations were determined using the BCA assay (Pierce™ BCA Protein Assay Kit; Thermo Fisher Scientific, A65453). Equal amounts of protein were resolved by SDS–PAGE and transferred onto 0.45-µm PVDF membranes (Invitrogen LC2005). Membranes were blocked for 1 hour at room temperature in 5% fat-free dry milk in TBS containing 0.1% Tween-20 (TBST) and incubated overnight at 4 °C with rabbit anti-TCRβ2 (ROX13; 0.001 µg/µL), mouse anti-TCRβ2 (MOX13; 0.001 µg/µL), or rabbit anti-β-tubulin (CST 2128T; 1:1000). After TBST washes, membranes were incubated for 1 hour at room temperature with HRP-conjugated anti-rabbit or anti-mouse secondary antibodies (Jackson ImmunoResearch). Signals were developed using SuperSignal™ West Atto Ultimate Sensitivity substrate (Thermo Fisher Scientific) and imaged with the ChemiDoc™ Touch Imaging System (Bio-Rad).

A pellet of each of the above cell lines was produced by centrifugation, clotted with human plasma (National Health Service Blood Transfusion Service, Cambridge, UK) and bovine thrombin (50 U/mL; BTUB291, Diagnostic Reagents Ltd, Thame, UK), marked, in a specific colour, with “Cancer Diagnostics Tissue Marking Dye” (CellPath, Newtown, Powys, UK) and processed to paraffin, as previously[1]. 3-micron paraffin sections were immunostained on charged microscope slides.

### FFPE Clinical Tissue Samples

Histological patient samples (Table 1) were obtained from the Cambridge University Hospitals NHS Foundation Trust Human Research Tissue Bank or “Research Histology” in the Department of Cellular Pathology, University Hospital Southampton NHS Foundation Trust, Southampton, UK, with full ethical approval (IRAS: 162057; PI: Professor E. Soilleux) [1].

**Table 1.**
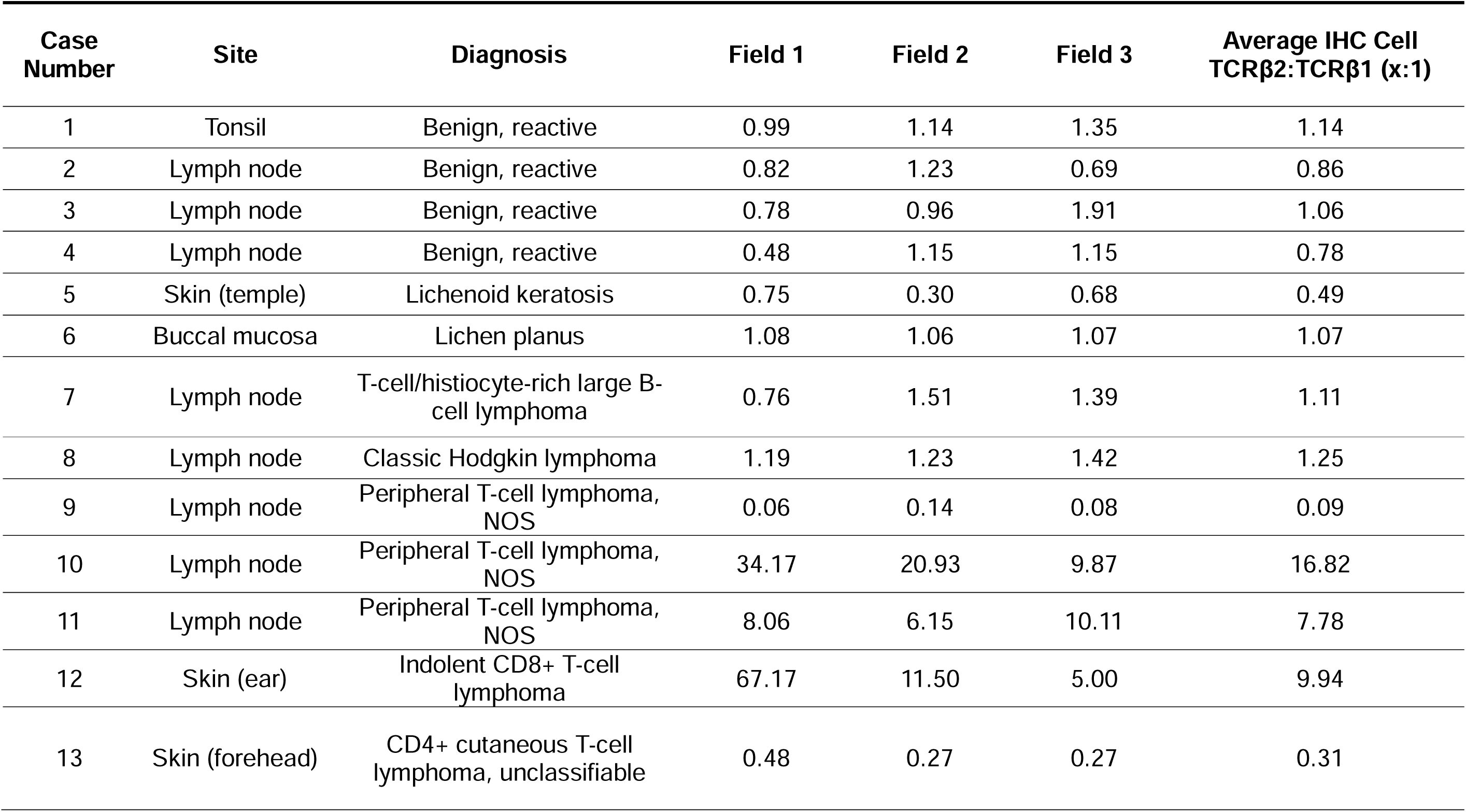
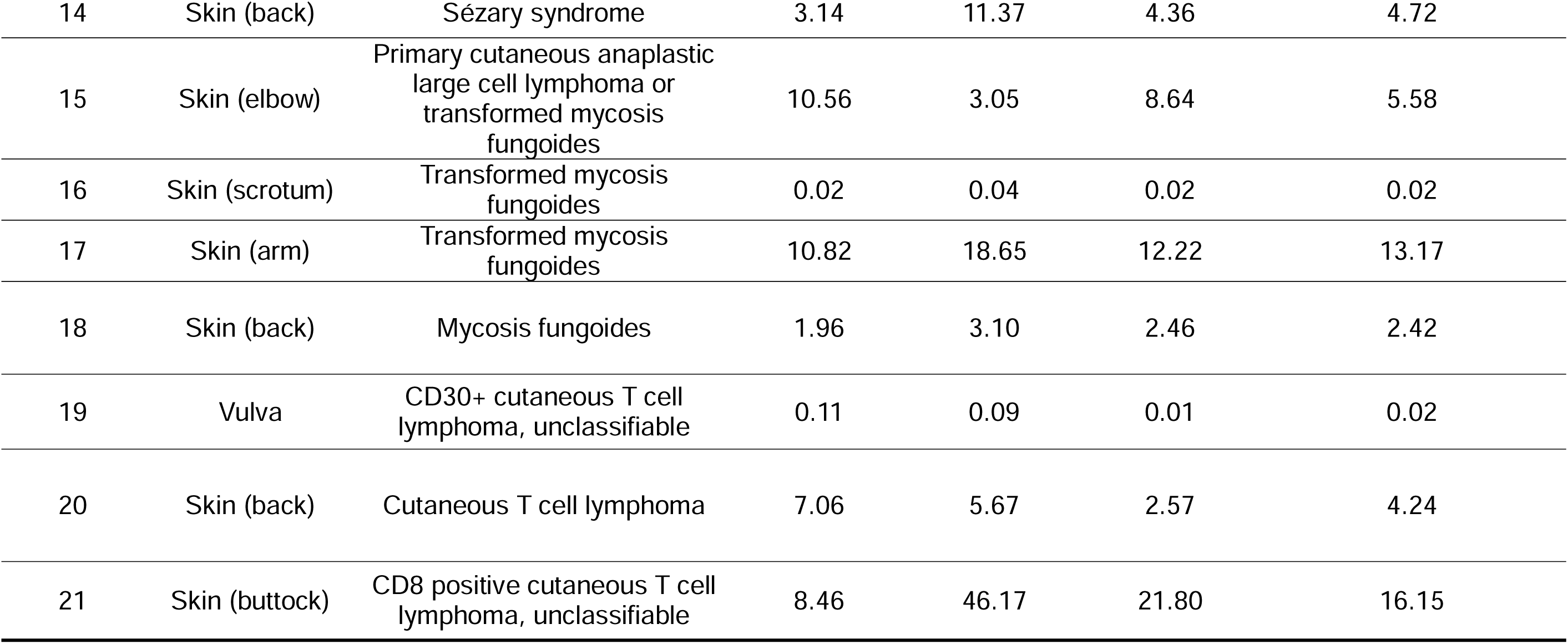
Analysis of TCRβ2+:TCRβ1+ cell immunostaining ratios in tissue sections containing benign and malignant T-cell populations, performed using the QuPath and StarDist software[11, 12], with human oversight. As benign T-cell populations showed a small excess of TCRβ2+ compared with TCRβ1+ cells, TCRβ2:TCRβ1 (rather than TCRβ1:TCRβ2) ratios were quantified. We have previously published quantitative real-time reverse transcription PCR (Q-PCR) and in situ hybridisation (ISH), determining the *TRBC2*:*TRBC1* ratios in these samples[1], all of which corroborate the immunostaining results in this table.

| Case Number | Site | Diagnosis | Field 1 | Field 2 | Field 3 | Average IHC Cell TCRβ2:TCRβ1 (x:1) |
| --- | --- | --- | --- | --- | --- | --- |
| 1 | Tonsil | Benign, reactive | 0.99 | 1.14 | 1.35 | 1.14 |
| 2 | Lymph node | Benign, reactive | 0.82 | 1.23 | 0.69 | 0.86 |
| 3 | Lymph node | Benign, reactive | 0.78 | 0.96 | 1.91 | 1.06 |
| 4 | Lymph node | Benign, reactive | 0.48 | 1.15 | 1.15 | 0.78 |
| 5 | Skin (temple) | Lichenoid keratosis | 0.75 | 0.30 | 0.68 | 0.49 |
| 6 | Buccal mucosa | Lichen planus | 1.08 | 1.06 | 1.07 | 1.07 |
| 7 | Lymph node | T-cell/histiocyte-rich large B-cell lymphoma | 0.76 | 1.51 | 1.39 | 1.11 |
| 8 | Lymph node | Classic Hodgkin lymphoma | 1.19 | 1.23 | 1.42 | 1.25 |
| 9 | Lymph node | Peripheral T-cell lymphoma, NOS | 0.06 | 0.14 | 0.08 | 0.09 |
| 10 | Lymph node | Peripheral T-cell lymphoma, NOS | 34.17 | 20.93 | 9.87 | 16.82 |
| 11 | Lymph node | Peripheral T-cell lymphoma, NOS | 8.06 | 6.15 | 10.11 | 7.78 |
| 12 | Skin (ear) | Indolent CD8+ T-cell lymphoma | 67.17 | 11.50 | 5.00 | 9.94 |
| 13 | Skin (forehead) | CD4+ cutaneous T-cell lymphoma, unclassifiable | 0.48 | 0.27 | 0.27 | 0.31 |
| 14 | Skin (back) | Sézary syndrome | 3.14 | 11.37 | 4.36 | 4.72 |
| 15 | Skin (elbow) | Primary cutaneous anaplastic large cell lymphoma or transformed mycosis fungoides | 10.56 | 3.05 | 8.64 | 5.58 |
| 16 | Skin (scrotum) | Transformed mycosis fungoides | 0.02 | 0.04 | 0.02 | 0.02 |
| 17 | Skin (arm) | Transformed mycosis fungoides | 10.82 | 18.65 | 12.22 | 13.17 |
| 18 | Skin (back) | Mycosis fungoides | 1.96 | 3.10 | 2.46 | 2.42 |
| 19 | Vulva | CD30+ cutaneous T cell lymphoma, unclassifiable | 0.11 | 0.09 | 0.01 | 0.02 |
| 20 | Skin (back) | Cutaneous T cell lymphoma | 7.06 | 5.67 | 2.57 | 4.24 |
| 21 | Skin (buttock) | CD8 positive cutaneous T cell lymphoma, unclassifiable | 8.46 | 46.17 | 21.80 | 16.15 |

### Immunostaining

Chromogenic immunohistochemical staining of cell pellets and patient samples was performed on a Leica Bond RX or Roche Ventana Discovery automated immunostainer, using rabbit IgG monoclonal antibodies against TCRβ1 (ROX7, 0.4 µg/mL) and TCRβ2 (ROX13, 2 µg/mL) or mouse IgG monoclonal antibodies against TCRβ2 (MOX13, 2 µg/mL). Antibodies were incubated for 60 minutes following heat-induced epitope retrieval for 60 minutes in alkaline buffer (Epitope Retrieval Solution 2; Cat. No. AR9640, Leica Biosystems, Newcastle-upon-Tyne, UK) or CC2 (ROX7) or CC1 (ROX13) (Cat. No. 950-224/ 950-223, Roche Diagnostics, Burgess Hill, UK). Detection was performed using the Polymer Refine Detection System (Cat. No. DS9800, Leica Biosystems, Newcastle-upon-Tyne, UK) or Optiview Detection System (Cat. No. 760-099, Roche Diagnostics, Burgess Hill, UK).

For double immunohistochemical staining, sequential staining was performed using antibodies ROX7, ROX13 or MOX13 as above, detected with the Polymer Refine Detection System (brown diaminobenzidine (DAB) chromogen), followed by staining with a second primary mouse or rabbit antibody (ROX13, MOX13, anti-CD4 (clone 4B12) or anti-CD8 (clone 4B11) (Leica, Newcastle, UK)), detected with the Bond Intense R Detection System (Cat. No. DS9263, Leica Biosystems, Newcastle-upon-Tyne, UK) (red chromogen).

Images were acquired at 100x and 400x magnification using an Infinity 2 camera (Lumenera, Ottawa, ON, Canada) mounted on a BX53 microscope (Olympus, Tokyo, Japan).

### Analysis of Histological Results

Three non-overlapping fields of serial sections immunostained with anti-TCRβ1 and anti-TCRβ2 antibodies were photographed at 400x, and the photographs used for quantitative analysis of immunostaining. Automated cell counting was performed in QuPath, using a pre-trained StarDist model, under human supervision[11, 12]. For each field, pixel size was adjusted to 0.4μm x 0.4μm and stain vectors were estimated from a representative region. Cells were classified based on mean DAB immunostaining intensity (strong/ weak) and size (large/ small). Positive cells of all sizes and intensities were counted. Counts were manually validated by three observers (blinded to diagnosis), including a practising haematopathologist (EJS).

## Results

### Western Blotting and immunostaining of FFPE pellets of immortalised cell lines

Western blotting and immunostaining of FFPE pellets of immortalised cell lines demonstrated the TCRβ2 specificity of our OX13 monoclonal antibodies, in both the original rabbit (ROX13) and chimeric mouse (MOX13) form (Figure 1). These anti-TCRβ2 antibodies gave strong and highly specific immunostaining at a much lower concentration than our previously published ROX11 anti-TCRβ2 antibody[1]. We confirmed the previously demonstrated TCRβ1-specificity of the rabbit monoclonal antibody, ROX7[1].

**Figure 1.**
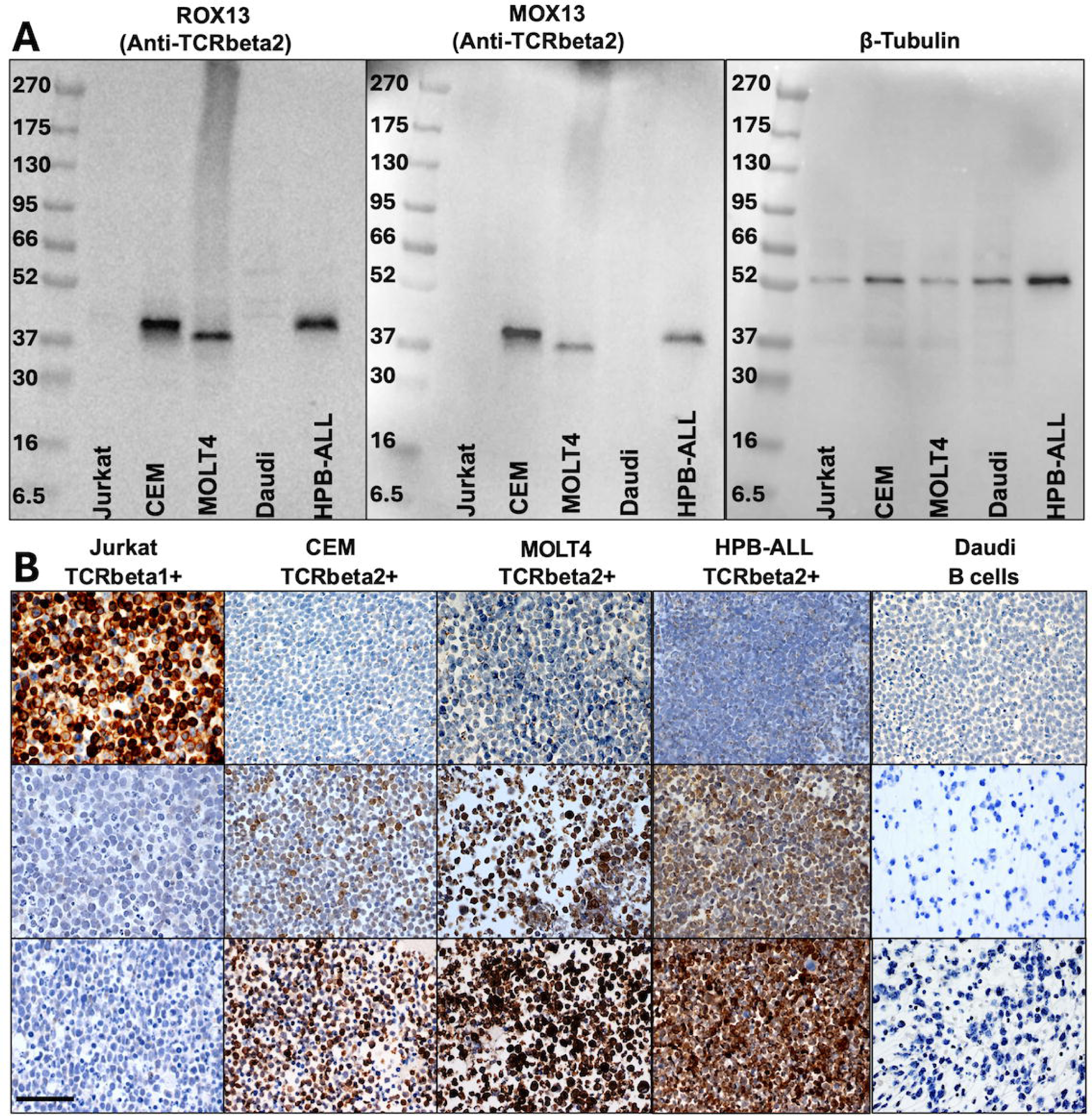
Validation of TCRβ isotype-specific monoclonal antibodies by Western blotting and immunohistochemistry. (A) Western blot demonstrating the specificity of TCRβ2-specific monoclonal antibodies ROX13 (rabbit) and MOX13 (mouse). Antibodies were validated using Jurkat (TCRβ1⁺), CEM (TCRβ2⁺), MOLT4 (TCRβ2⁺), Daudi B (TCRβ-negative), and HPB-ALL (TCRβ2⁺) cell lines. Monoclonal rabbit anti-β-tubulin is used as a protein loading control. (B) Chromogenic immunostaining of FFPE cell pellets from Jurkat, CEM, MOLT4, Daudi, and HPB-ALL cell lines. The high specificity of ROX7 (TCRβ1-specific), ROX13 (TCRβ2-specific), and MOX13 (TCRβ2-specific) antibodies is demonstrated. Target antigens are visualised as brown staining using diaminobenzidine (DAB), with nuclei counterstained blue by haematoxylin. Scale bar in bottom left panel represents 50 microns and pertain to all panels. Compared with our previously published ROX11 anti-TCRβ2 monoclonal antibody[1], the ROX13 and MOX13 antibodies give stronger positive immunostaining, with minimal non-specific staining, at a much lower concentration than ROX11.

### Utility of the anti-TCRβ1/anti-TCRβ2 pair in separating benign and malignant (lymphomatous) T-cell populations

In the benign T-cell populations in FFPE lymph node and tonsil tissue shown in Figure 2A, there are roughly equal numbers of TCRβ1+ (ROX7+) and TCRβ2+ (ROX13+) T cells, both in the paracortex of lymph nodes the T-zones of tonsils, as well as among the follicular helper T cells of B-cell follicles. Examples of T-cell lymphomas with clear TCRβ1 or TCRβ2 restriction are shown in Figure 2B. Compared with clonality testing, our immunostaining has the advantages of allowing assessment of both the cellular morphology and distribution (lesional architecture) of TCRβ1/2+ T cell infiltrates. Case 9 (Figure 2) shows TCRβ1-restriction of the large lymphoid cells, while case 20 (Figure 3) demonstrates clear TCRβ2-restriction of its epidermotropic T-cell population.

**Figure 2.**
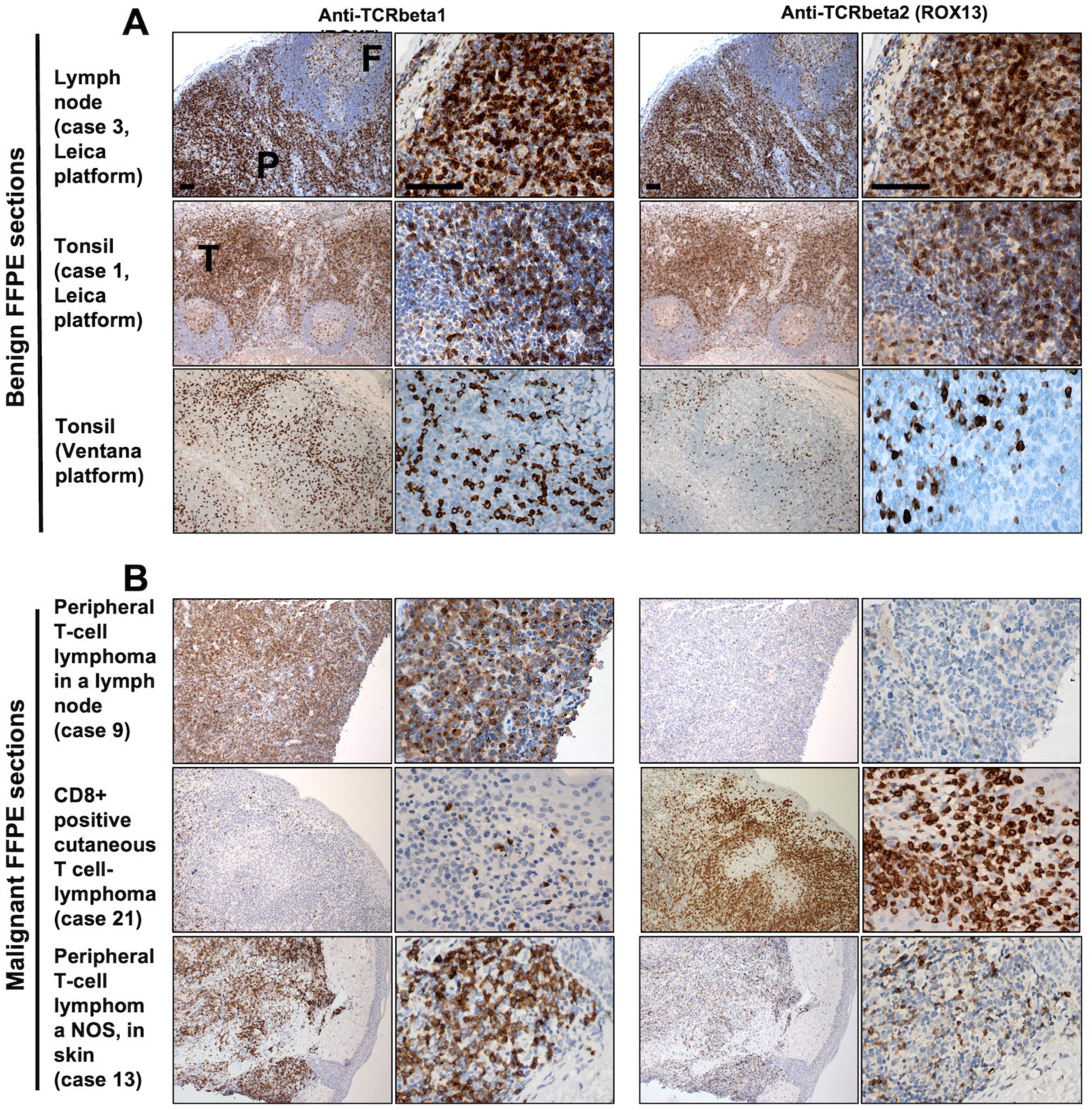
Validation of anti-TCRβ1/2 monoclonal antibodies for immunohistochemistry on FFPE samples. Representative images of benign (A) and malignant (B) tissue sections immunostained with ROX7 (TCRβ1 specific; left panels) and ROX13 (TCRβ2 specific; right panels). Benign sections show positive immunostaining (brown) of T cells in the lymph node paracortex (P), tonsillar T zone (T) and among the scattered T-cell population in B-cell follicles (example labelled F). Comparable numbers of cells are stained positively with each antibody, on both the Leica (top two rows) and Ventana immunostaining platforms (lower row). In B, the upper and lower panels represent TCRβ1-restricted lymphomas, whereas the middle panel represents a TCRβ2-restricted lymphoma. Compared with clonality testing, our TCRβ1/2 immunostaining has the advantage of allowing assessment of the distribution (lesional architecture) of TCRβ1/2+ T cell infiltrates. Furthermore, TCRβ-restriction can be assessed in cell populations with specific morphology. Case 9, for example, shows that the large lymphoid cell population is TCRβ1-restricted. Case numbers refer to the cases described in Table 1. Scale bars in top row panels represent 50 microns and pertain to all panels in the same column.

**Figure 3.**
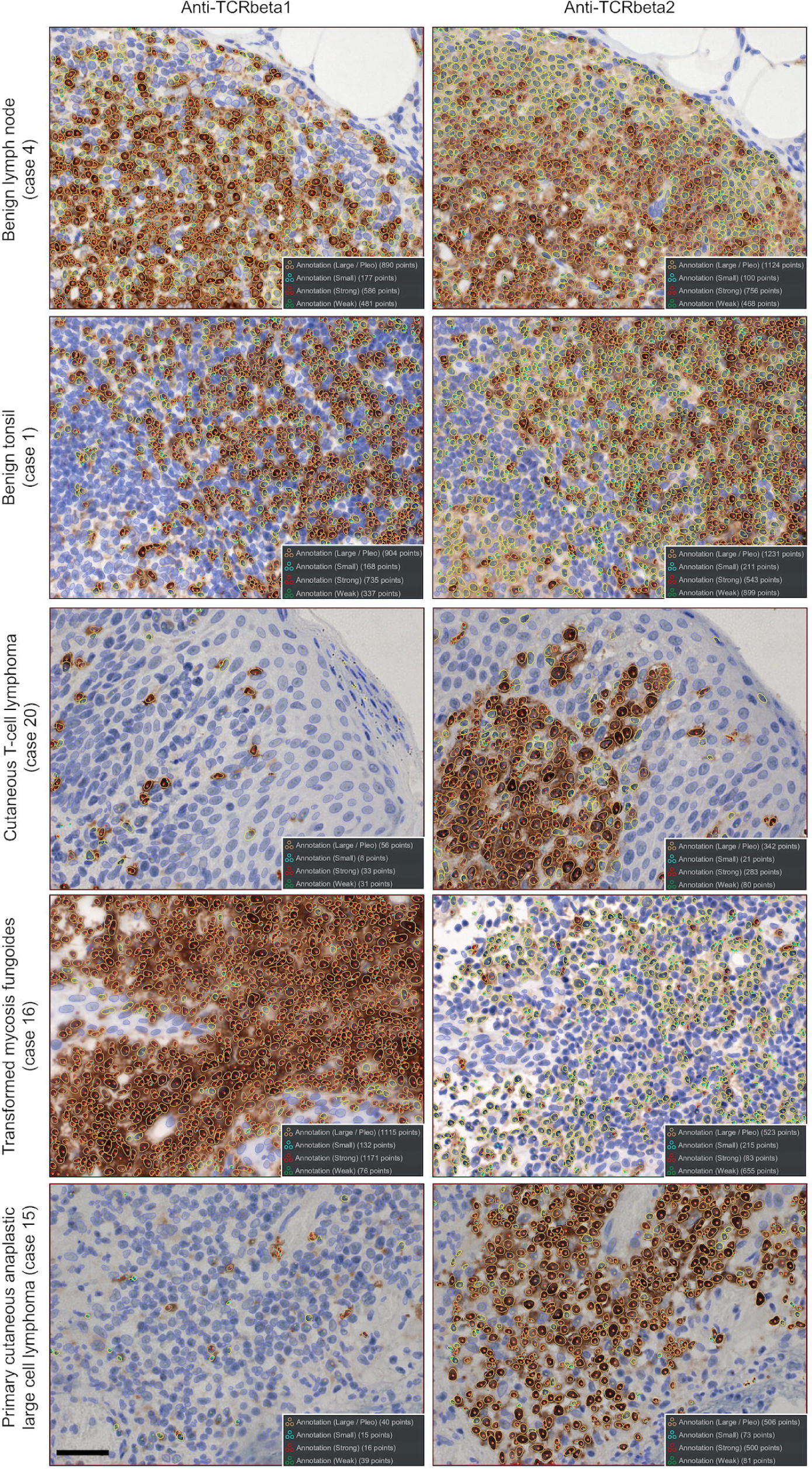
Analysis of tissue sections containing benign and lymphomatous T-cell populations using an automated cell counting workflow. Tissue sections were immunostained (brown) with anti-TCRβ1 (ROX7) and anti-TCRβ2 (ROX13) antibodies with a blue haematoxylin nuclear counterstain. Cell detection was performed using StarDist. Cells were categorised based on staining intensity and size, then quantified. Annotated outlines around cells indicate diaminobenzidine (DAB) immunostaining intensity: orange (strongly stained), yellow (weakly stained) and blue (negative). Dots overlying the nuclei indicate exactly which nuclei were counted, and their size and diaminobenzidine (DAB) immunostaining intensity (key in bottom right of each panel). The corresponding case number of each image and diagnosis are indicated to the left of each row. Scale bar in bottom left-hand panel represents 50 microns and pertains to all panels. Case 20, a cutaneous T-cell lymphoma with morphological features of mycosis fungoides, demonstrates clear TCRβ2-restriction of its epidermotropic T-cell population. This underlines the advantage of our immunohistochemical surrogate for clonality testing over conventional DNA-based clonality analysis[3, 7], because our immunohistochemical approach permits simultaneous assessment of cellular morphology and immunophenotype[1].

### Application of an automated cell counting method (with human oversight) to the immunostained sections

While TCRβ restriction, a surrogate for T-cell monoclonality, is obvious in many T-cell lymphomas, the admixture of benign T cells, which constitute a roughly equal TCRβ1+/2+ mixture, may confound this assessment. In this situation, counting of cells may be necessary, and can prove laborious. We therefore tested whether user-friendly free software could perform this task. A combination of the QuPath and StarDist software, with human oversight, was used to quantify positively immunostained cells in 3 paired (TCRβ1/2) serial section images (Figure 3)[11, 12]. The mean TCRβ2:TCRβ1 ratios in benign T-cell populations were between 0.49:1 and 1.25:1, with the ratios in all lymphoma samples falling outside this (Table 1). We thus demonstrated that an automated cell counting method (with human oversight) could be applied to TCRβ1/2-immunostained FFPE sections.

### Anti-TCRβ1/2 double immunostaining could facilitate diagnosis

The chimeric mouse version of our anti-TCRβ2 antibody (MOX13) gave very similar staining to that of the rabbit parent antibody (ROX13). We tested both these anti-TCRβ2 antibodies, together with our anti-TCRβ1 (ROX7) antibody in double immunostaining on the Leica Bond Rx platform. Similar numbers of TCRβ1+ and TCRβ2+ cells were seen in the immunostained sections as expected (Figure 4A). Double immunostaining of T-cell lymphomas with the rabbit anti-TCRβ1/2 antibody pair permitted visualization of TCRβ restriction in two examples of cutaneous T-cell lymphoma (Figure 4B). These results show that double immunostaining could be used diagnostically.

**Figure 4.**
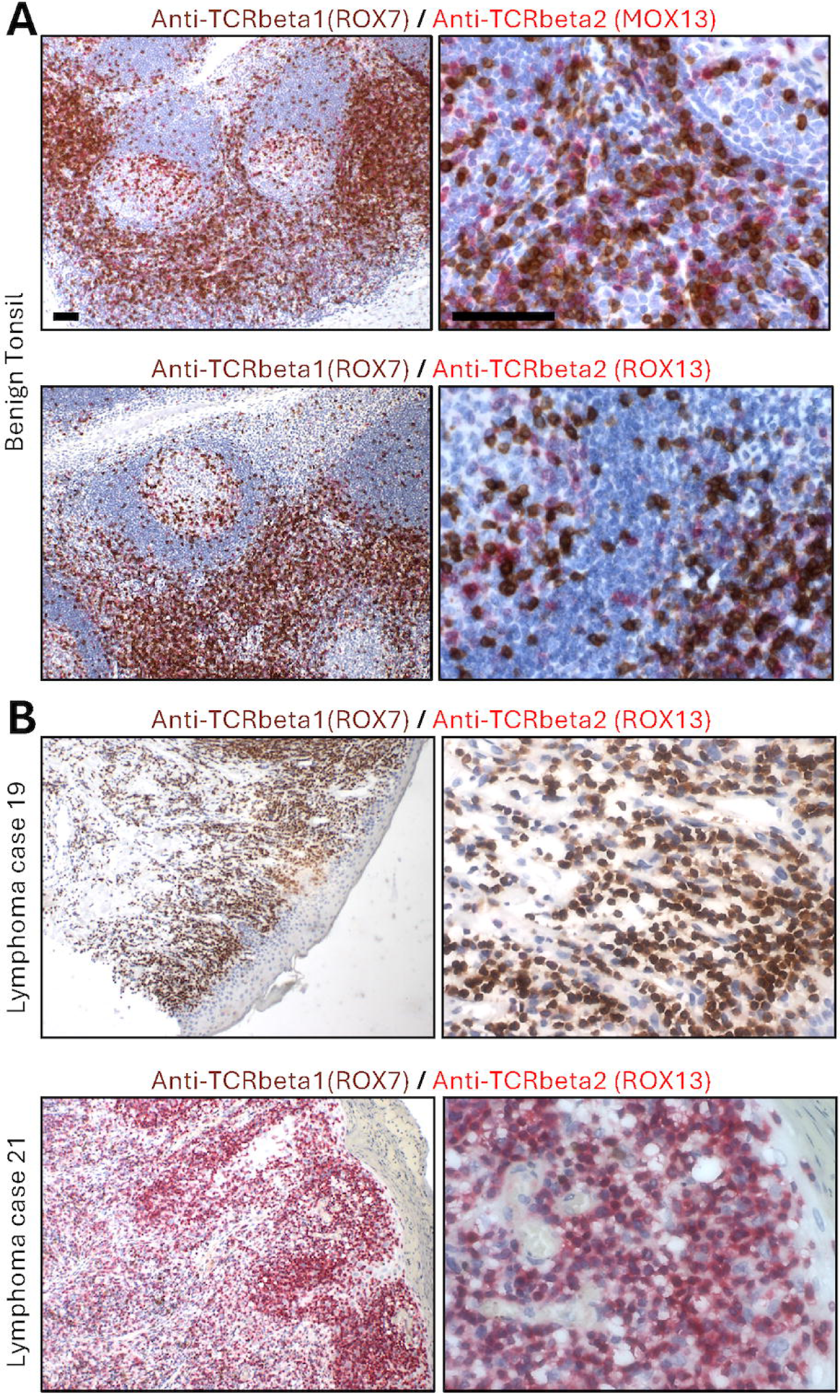
Validation of anti-TCRβ1/2 monoclonal antibodies for double immunostaining with rabbit anti-TCRβ1 (brown) and mouse/ rabbit anti-TCRβ2 (red) antibodies, on benign (A) and malignant (B) T-cell populations. (A) Tonsil (upper panel) shows roughly equal numbers of TCRβ1+ (brown) and TCRβ2+ (red, detected with mouse antibody). (B) TCRβ1-restricted CD30+ cutaneous T-cell lymphoma, unclassifiable (case 19; upper panel) and TCRβ2-restricted cutaneous lymphoma (case 21; lower panel) immunostained. Panels in B show immunostaining with rabbit anti-TCRβ1 (brown) and mouse anti-TCRβ2 (red) antibodies. Scale bars in top row panels represent 50 microns and pertain to all panels in the same column.

### Potential utility of anti-TCRβ1/2 double immunostaining with CD4 or CD8

In lymph nodes, around two-thirds of benign, mature T cells are CD4+ CD8-, while almost all of the remainder are CD4-CD8+[13]. T-cell lymphomas are often CD4+, less commonly CD8+, and occasionally CD4-CD8-[2]. Assay sensitivity for TCRβ restriction in suspected malignant T-cell populations could be further increased by assessing TCRβ1/2 expression in individual T-cell populations separated on the basis of their CD4 and CD8 expression. As proof-of-concept, we show examples of TCRβ1/2 double immunostaining, together anti-CD4/CD8. Figure 5 shows two cutaneous T-cell lymphomas, one weakly CD4+, TCRβ1-restricted and one CD8+, TCRβ2-restricted. A larger study will be necessary for full assessment of the utility of anti-TCRβ1/2 double immunostaining with anti-CD4/ CD8.

**Figure 5.**
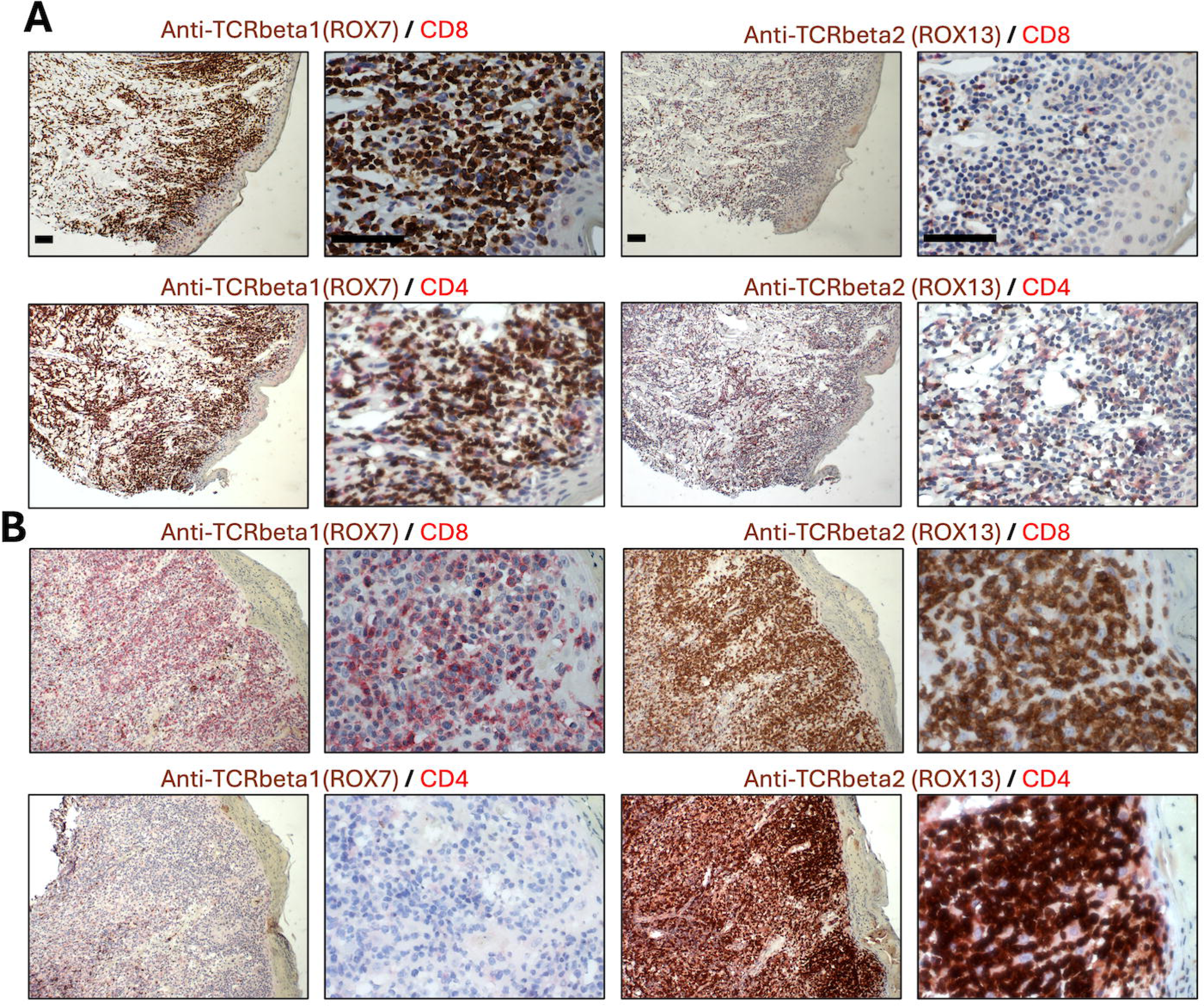
Validation of anti-TCRβ1/2 monoclonal antibodies for distinguishing tumour-infiltrating from malignant T-cell populations. Double immunostaining of cutaneous lymphomas showing TCRβ1-restriction (case 19; panels in A) or TCRβ2-restriction (case 21; panels in B), performed using anti-TCRβ1 (ROX7; brown) or anti-TCRβ2 (ROX13; brown) antibodies in combination with anti-CD8 (upper panels; red) or anti-CD4 (lower panels; red) antibodies. Immunostaining demonstrates that the TCRβ1-restricted lymphoma cells are CD4+, whereas the TCRβ2-restricted lymphoma cells are CD8+. Scale bars in top row panels represent 50 microns and pertain to all panels in the same column.

## Discussion

Here we validate a pair of antibodies for single and double immunostaining that could substantially improve and expedite the diagnosis of T-cell lymphoma, decreasing time-to-diagnosis by around two weeks. The diagnosis of T-cell lymphoma relies initially on immunostaining multiple serial sections with immunophenotypic markers, including CD2, CD3, CD4, CD5, CD7, CD8, CD25, CD30. Adding our antibodies to the diagnostic armoury requires just two additional serial sections, although double or multiplexed immunostaining with our antibodies and CD4 or CD8 could also be undertaken, if clinically useful. Immunostaining with our antibodies, unlike clonality testing, can be undertaken in non-specialist centres. While some guidelines (e.g., in the UK) mandate only diagnosing lymphomas in specialist haemato-oncology diagnostic units[14], the antibodies could be very valuable in non-specialist centres, to triage specimens for referral to specialist centres.

TCRβ-restriction was obvious in many of our lymphoma cases, but, in some lymphomas, a significant tumour-infiltrating benign T-cell population confounds analysis. We show that such cases are amenable to automated cell counting. This approach permitted calculation of a benign/ malignant TCRβ2:TCRβ1 ratio cut-off of 0.49 – 1.25:1. This ratio will be further refined with a subsequent larger study of T-cell populations in various non-lymphomatous pathologies, across a range of body sites and patient ages and through double immunostaining.

Double immunostaining was highly successful with our TCRβ1 and TCRβ2 rabbit antibodies. Our chimeric mouse anti-TCRβ2 antibody, which performs very similarly to the parent rabbit anti-TCRβ2 antibody, can facilitate double immunostaining in stings in which antibodies of two different animal species are required. Similarly, we demonstrated the potential utility of double immunostaining with each of our TCRβ1/2 antibodies and anti-CD4/ CD8 antibodies. However, co-localisation of the chromogens can make interpretation difficult, as CD4, CD8 and TCRβ1/2 are all found in a membranous/ cytoplasmic location. Instead of looking for co-localisation, CD4 or CD8 immunostaining might be better used to identify the T-cell population in which TCRβ1/2 assessment is not required (i.e., a T-cell population to be ignored). Using this strategy, double immunostaining with anti-CD8 may be more useful than anti-CD4, because fewer T-cell lymphomas are CD8+. Larger studies involving multiple pathologists are required to determine the utility of these double immunostaining approaches in clinical diagnosis.

Although TCRβ1/2 antibody pairs have been used in flow cytometry[15], this and our previously published antibody pairing are the only examples, to our knowledge, of an anti-TCRβ1/ anti-TCRβ2-specific monoclonal antibody pair that works on FFPE material, making it suitable for solid haematopathological specimens. This approach, like kappa and lambda detection, for B-cell lymphoma and myeloma, will expedite and improve the triage and diagnosis of T-cell lymphomas, particularly cutaneous lymphomas and those with small/medium cellular morphology. We anticipate wide adoption in histopathology and haematopathology departments internationally.

## Data Availability

All data produced in the present work are contained in the manuscript.

## Acknowledgments

We are grateful to the Cambridge University Hospitals NHS Foundation Trust Human Tissue Research Biobank and to the Histology Research team at the University Hospital of Southampton.

## Competing Interests

E.J.S., S.C.E., and G.O. are co-inventors on patents granted and/or filed related to the diagnosis and treatment of T-cell lymphoma.

## Funding

This work was funded by Cancer Research UK, reference EDDPJT-Nov24/100031. GO is supported by the Medical Research Council. Original antibody discovery was supported by Oxford University Innovation Challenge Seed Fund.

## Notes

### Author Declarations

This study was undertaken with full ethical approval (IRAS: 162057; PI: Professor E. Soilleux) from Oxfordshire Research Ethics Committee A (now South Central - Oxford A)

